# The time scale of asymptomatic transmission affects estimates of epidemic potential in the COVID-19 outbreak

**DOI:** 10.1101/2020.03.09.20033514

**Authors:** Sang Woo Park, Daniel M. Cornforth, Jonathan Dushoff, Joshua S. Weitz

**Author notes:** Electronic address; URL: https://mac-theobio.github.io/dushoff.html. Electronic address; URL: http://ecotheory.biology.gatech.edu.

## Abstract

The role of asymptomatic carriers in transmission poses challenges for control of the COVID-19 pandemic. Study of asymptomatic transmission and implications for surveillance and disease burden are ongoing, but there has been little study of the implications of asymptomatic transmission on dynamics of disease. We use a mathematical framework to evaluate expected effects of asymptomatic transmission on the basic reproduction number *ℛ*_0_ (i.e., the expected number of secondary cases generated by an average primary case in a fully susceptible population) and the fraction of new secondary cases attributable to asymptomatic individuals. If the generation-interval distribution of asymptomatic transmission differs from that of symptomatic transmission, then estimates of the basic reproduction number which do not explicitly account for asymptomatic cases may be systematically biased. Specifically, if asymptomatic cases have a shorter generation interval than symptomatic cases, *ℛ*_0_ will be over-estimated, and if they have a longer generation interval, *ℛ*_0_ will be under-estimated. Estimates of the realized proportion of asymptomatic transmission during the exponential phase also depend on asymptomatic generation intervals. Our analysis shows that understanding the temporal course of asymptomatic transmission can be important for assessing the importance of this route of transmission, and for disease dynamics. This provides an additional motivation for investigating both the importance and relative duration of asymptomatic transmission.

## I. INTRODUCTION

In an epidemic, symptomatic cases are the predominant focus of treatment and usually represent the bulk of reported cases. However, infected individuals who are asymptomatic yet infectious can be a critical factor in the spread of some pathogens [1]. Asymptomatic individuals are hard to trace, unlikely to self-isolate, and are likely to retain normal social and travel patterns [2].

There is significant ongoing interest in asymptomatic infections in COVID-19 [3–5] and their transmission potential [6] for two major reasons. First, the proportion of infections that are asymptomatic (see [7]) is critical to attempts to estimate the likely burden of severe outcomes (including mortality [8]) when the virus spreads through a population. Second, understanding the possible role of *transmission* by asymptomatic individuals is crucial to planning surveillance and control efforts [1]. Given that 86% of the cases were undocumented (i.e., mildly symptomatic or asymptomatic) in Wuhan prior to travel restrictions and may account for 79% of infection in severe, symptomatic cases [9], asymptomatic cases are also likely to play an important role in the transmission of COVID-19.

Here, we focus on a third effect. If asymptomatic cases are important for transmission, they also have the potential to affect estimates of key parameters of disease spread such as the basic reproduction number *ℛ*_0_ (i.e., the expected number of secondary cases generated by an average primary case in a fully susceptible population [10]). Thus, we investigate the relationship between individual-level features of asymptomatic cases (e.g., the probability that an infection is asymptomatic, asymptomatic case duration, transmission by asymptomatic individuals) to dynamics at the population scale.

## II. METHODS

We model viral spread using a renewal-equation framework [11], which allows us to model the current incidence of infected individuals (i.e., the rate at which new infections occur in the population) as a function of previous incidence and how infectiousness of an infected individual varies over the course of their infection. We divide incidence *i* into two categories –*i*_*a*_ and *i*_*s*_ – corresponding to incidence of asymptomatic and symptomatic cases, respectively. Newly infected individuals that are either asymptomatically or symptomatically infected can transmit the disease to others, but they may differ in their intrinsic reproduction numbers, *ℛ*_*a*_ and *ℛ*_*s*_, respectively, as well as intrinsic generation-interval distributions [12], *g*_*a*_(*τ*) and *g*_*s*_(*τ*). Generation intervals, which are defined as the time between when an individual is infected and when that individual infects another person [13], shape the relationship between the epidemic growth rate *r* and the reproduction number [14]. The differences in the generation-interval distributions between asymptomatic and symptomatic cases can be caused by the differences in the natural history of infection irrespective of their transmissibility: Individuals with asymptomatic infections may recover faster and have short generation intervals, or have persistent infection and long generation intervals (cf. [15]).

Neglecting births and loss of immunity on the time scale of the outbreak, the dynamics of susceptibles and incidence are (see Table S1 for parameter definitions):

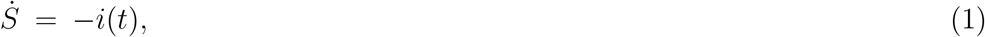

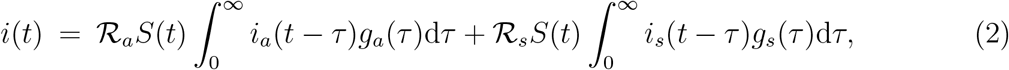

where *i*(*t*) = *i*_*a*_(*t*) + *i*_*s*_(*t*). The basic reproduction number of this system is:

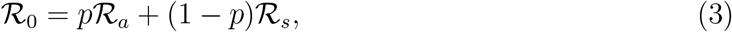

where *p* is the proportion of *incident cases* that are asymptomatic: *i*_*a*_(*t*) = *pi*(*t*). The corresponding intrinsic generation-interval distribution of an average infected individual is given by:

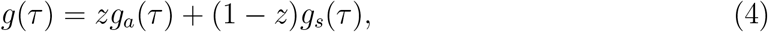

where we define the “intrinsic” proportion of asymptomatic transmission *z* as the relative contribution of asymptomatic cases to the basic reproduction number:

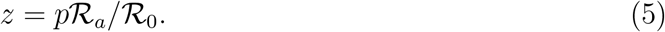

Note that the intrinsic proportion of symptomatic transmission satisfies

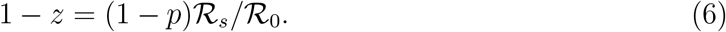

Yet, this information is not sufficient to disentangle the role of asymptomatic cases, i.e., what fraction of secondary cases can be ascribed to *realized* transmission from asymptomatic cases vs. symptomatic cases?

The intrinsic proportion of asymptomatic transmission *z* is a useful benchmark, but does not necessarily reflect the realized proportion of asymptomatic transmission, unless both types of infection have the same generation-interval distribution. The *realized* proportion of asymptomatic transmission, *q* at time *t* is given by:

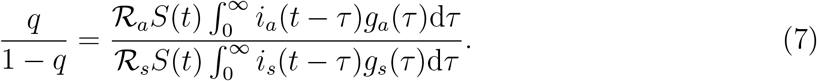

During the period of exponential growth, we assume *S* remains nearly constant, and *i*(*t*) is proportional to exp(*rt*); here, the observed exponential growth rate *r* is an average of the exponential growth rates we would observe if there were only asymptomatic (*p* = 1) or symptomatic (*p* = 0) cases. We then simplify by recalling that *i*_*a*_(*t*) = *pi*(*t*), *i*_*s*_(*t*) = (1*−p*)*i*(*t*) such that:

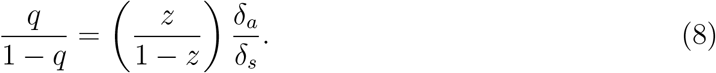

Here, *δ*_*c*_ for each of the two classes is the average “discount” of a new infection – the average relative contribution of a secondary infection to the epidemic, taking exponential growth into account:

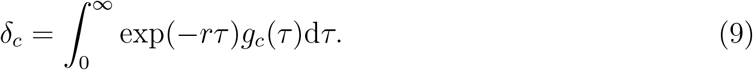

*δ*_*c*_ *<* 1 and grows smaller as the generation interval grows longer. Thus, the realized proportion of asymptomatic infections will be increased (resp., decreased) if transmission is relatively faster (slower) along the asymptomatic route. The discount *δ* also depends on the relative variation in the generation-interval distribution, the “dispersion”: More variation in generation intervals leads to more opportunities for fast spread and thus to higher values of *δ* (similar to shorter average generation intervals).

To estimate the effects of assumptions about asymptomatic transmission on the inferred importance of asymptomatic transmission and estimates of the basic reproduction number *ℛ*_0_, we parameterize the generation interval distributions of asymptomatic and symptomatic cases based on their means, 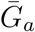 and 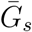, and dispersions, *κ*_*a*_ and *κ*_*s*_. We assume that generation intervals are gamma distributed, and we set the dispersion to be equal to the squared coefficient of variation (the reciprocal of the gamma shape parameter, see Supplementary Materials). We assume that epidemic growth rate *r* and the generation-interval distribution of symptomatic case are known, using parameter values that are consistent with earlier COVID-19 models [16]: 1*/r* = 7 days, 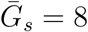 days, and *κ*_*s*_ = 0.5. We infer values of *q* using

Eq. (8) and *ℛ*_0_ using the Euler-Lotka equation [17]:

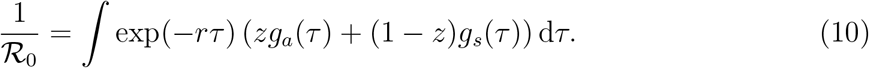

We compare this with the naive estimate of the basic reproduction number that assumes that the generation-interval distributions of the asymptomatic and symptomatic cases are identical:

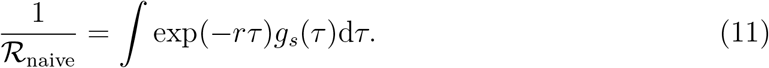

In Supplementary Materials, we also use an ordinary differential equation model (SEIR model) including both asymptomatic and symptomatic cases to give a concrete example of how differences in generation intervals affect both *q* and estimates of *ℛ*_0_.

## III. RESULTS

We explore the effects of different assumptions about speed and effectiveness of asymptomatic transmission on the importance of asymptomatic transmission and estimates of the basic reproduction number *ℛ*_0_, using a gamma assumption (see Methods). Across the range of parameters we explore, the intrinsic proportion of asymptomatic transmission *z* is similar to the realized proportion *q* (Figure 1A). As the relative mean generation interval of asymptomatic transmission, 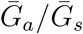, increases, *q* decreases because symptomatic cases are more likely to have short generation intervals, which drive the spread during the growth phase (Figure 1A). In Figure S1, we present the same figure but showing differences between the realized and the intrinsic proportion of asymptomatic transmission, *q − z*.

**FIG. 1:**
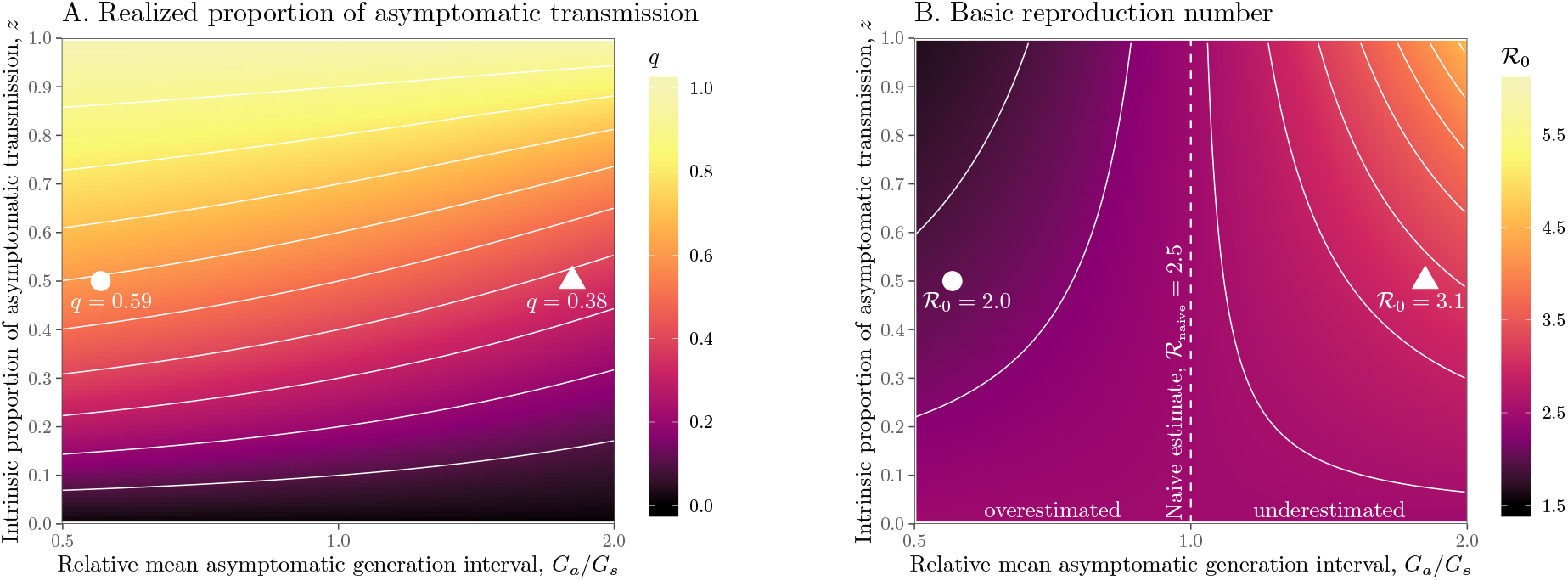
Effects of intrinsic proportion of asymptomatic transmission on the realized proportion of asymptomatic transmission and basic reproduction number, given variation in the mean generation interval of asymptomatic cases. (A) Increasing the speed of asymptomatic transmission (shorter generation intervals) increases the realized proportion of asymptomatic transmission, *q*. Increasing the speed of asymptomatic transmission (shorter generation intervals) decreases the basic reproduction number *ℛ*_0_. When 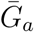is smaller (larger) than 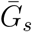, estimates based on the observed generation distribution for symptomatic cases (*ℛ*_0_ = 2.5; dashed line) are expected to over- (under-) estimate the true *ℛ*_0_. For both panels, the circle denotes *z* = 0.5 and 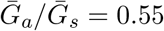 whereas the triangle denotes *z* = 0.5 and 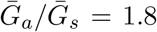. Solid lines show contours for *q* and *ℛ*_0_ values. The dashed line represents the naive estimate that assumes 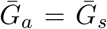. Here, we assume 1*/r* = 7 days, 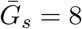 days, and *κ*_*s*_ = *κ*_*a*_ = 0.5.

Figure 1B shows the effect of different assumptions about the generation interval of asymptomatic cases, 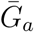, on the estimated basic reproduction number *ℛ*_0_. When 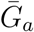 is long compared to 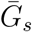, then we are effectively assuming a longer mean for the overall generation interval. This assumption leads to a larger estimate of *ℛ*_0_ for a fixed value of *r* (see [18]). Conversely, when 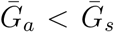, generation intervals are shorter, leading to lower estimates of the epidemic strength *ℛ*_0_. Both of these effects are stronger when the intrinsic proportion of asymptomatic transmission *z* increases (and disappear as *z→*0). Therefore, when *ℛ*_0_ is estimated without explicitly accounting for asymptomatic spread (white, dashed line in Figure 1B), it can be over- or underestimated depending on the relative duration of infection between symptomatic and asymptomatic individuals. The qualitative effects of *z* and 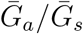 on *q* and *ℛ*_0_ remain robust when we assume narrower (*κ*_*s*_ = *κ*_*a*_ = 0.3; Figure S2) or wider (*κ*_*s*_ = *κ*_*a*_ = 0.8; Figure S3) generation intervals.

Relative generation-interval dispersion of asymptomatic cases *κ*_*a*_*/κ*_*s*_ have similar, but smaller, effects on *q* and *ℛ*_0_ (Figure S4). Since a wider generation-interval distribution has a higher proportion of early transmission than a narrow one, increasing the generation-interval dispersion has qualitatively similar effects on *q* and *ℛ*_0_ as decreasing the mean generation interval.

## IV. DISCUSSION

Much is still unknown about the time scale and effectiveness of asymptomatic transmission in COVID-19. Here we highlight the need to characterize the generation-interval distribution for asymptomatic transmission, and its consequences not only for contact tracing but for estimation of the basic reproduction number of the ongoing COVID-19 outbreak [16] and of the effective proportion of asymptomatic transmission during the exponential-growth phase. Our reproductive number findings fit into a broader framework linking epidemic speed, strength, and generation intervals – for a given observed speed increases in the mean generation interval imply larger reproduction number [14, 15, 18–20].

If asymptomatic infections are more persistent than symptomatic ones, the mean generation interval for COVID-19 could be longer than estimated from symptomatic cases alone possibly causing *ℛ*_0_ to be underestimated (Figure 1B). However, if asymptomatic cases tend to resolve quickly, then current estimates of *ℛ*_0_ may be over-estimates of the underlying strength (Figure 1B), and asymptomatic cases may be driving a larger fraction of secondary cases than we would expect without accounting for their differences (Figure 1A). The importance of these effects depends on the relative infectiousness of asymptomatic transmission as well as the proportion of incident cases that are asymptomatic (and therefore the intrinsic proportion of asymptomatic transmission *z*). The biases in the estimates of *ℛ*_0_ will necessarily bias estimates of the amount of intervention required to control the epidemic. Note that cases do not have to be completely asymptomatic for our qualitative results to apply. People with mild symptoms unlikely to be diagnosed in a particular time and place (sometimes referred to as subclinical cases) are expected to affect transmission patterns in the same way.

We focus here on the exponential phase, so it is worth noting that the realized proportion of asymptomatic transmission *q* is time-dependent, varying with dynamic changes in incidence and proportion susceptible. Future work might also consider the ways in which asymptomatic individuals can modulate the catalysis of epidemics in a networked metapopulation [21–23]. Characterizing the role of asymptomatic individuals in driving the persistence of the epidemic will be critical for assessing the post-pandemic outcome [24].

## Data Availability

All code is available at https://github.com/mac-theobio/coronavirus_asymptomatic.

## Acknowledgments

The authors thank John Glasser for comments and discussion on the manuscript, particularly on short notice. The authors thank multiple reviewers for their feedback. Research effort by JSW was enabled by support from grants from the Simons Foundation (SCOPE Award ID 329108), the Army Research Office (W911NF1910384), National Institutes of Health (1R01AI46592-01), and National Science Foundation (1806606 and 1829636). JD was supported, in part, by grants from the Canadian Institutes of Health Research.

## Data Availability

All code is available at https://github.com/mac-theobio/ coronavirus_asymptomatic.

## Supplementary Materials

### A gamma approximation to generation-interval distributions

Assuming that the intrinsic generation-interval distribution for each class (asymptomatic and symptomatic) follows a gamma distribution with mean 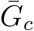 and dispersion *κ*_*c*_, the average discount of a new infection can be written as:

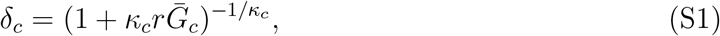

which shows that the average discount increases with smaller 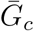 and with larger *κ*_*c*_. Then, the odds of the realized proportion of asymptomatic transmission can be written as:

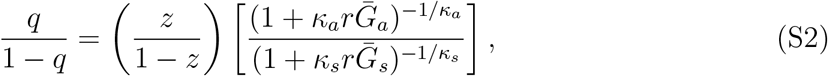

where 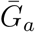 and 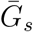 are the mean generation intervals for asymptomatic and symptomatic cases, and *κ*_*a*_ and *κ*_*s*_ are the generation-interval dispersions for asymptomatic and symptomatic cases. Finally, the basic reproduction number is calculated by using the Euler-Lotka equation:

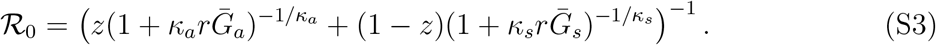

### A compartmental model for asymptomatic/symptomatic cases

Consider an SEIR model variant in which an infected individual can be either asymptomatic, *I*_*a*_, or symptomatic, *I*_*s*_. We note that *I*_*a*_ and *I*_*s*_ represent prevalence (i.e., the total number of currently infectious individuals) of asymptomatic and symptomatic individuals; these quantities are different from *i*_*s*_ and *i*_*s*_ that we present in the main text, which represent incidence (i.e., the rate at which new cases are generated) of asymptomatic and symptomatic individuals. While both cases can recover, we assume that only symptomatic cases can lead to fatalities, denoted by the *D* category. In total, the dynamics of susceptibles, exposed, infectious, recovered, and dead are:

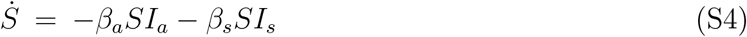

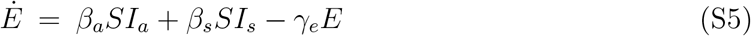

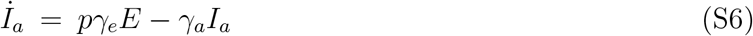

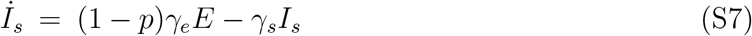

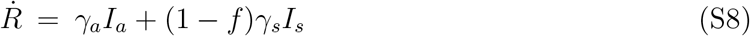

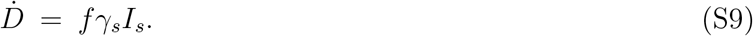

Here, *β*_*a*_ and *β*_*s*_ denote transmission rates, *γ*_*e*_ denotes the transition from exposed to infectious, *p* is the fraction of asymptomatic cases that are generated for each exposed individual, 1 *− p* is the fraction of symptomatic cases that are generated for each exposed individual, *γ*_*a*_ and *γ*_*s*_ denote recovery rates, and *f* denotes the case fatality ratio for symptomatic cases.

Given that the number of infected individuals increase exponentially at rate *r* initially, the equations for the infectious cases can be rewritten given the ansatz *E*(*t*) = *c*_*e*_*e*^*rt*^, *I*_*a*_(*t*) = *c*_*a*_*e*^*rt*^, *I*_*s*_(*t*) = *c*_*s*_*e*^*rt*^. Then, it follows that

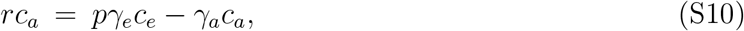

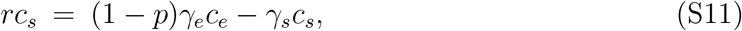

which implies that

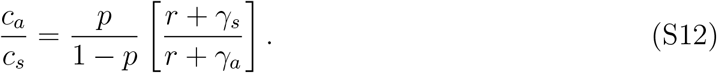

This shows that the prevalence of asymptomatic and symptomatic individuals is different from *p* and 1 *− p* because prevalence measures the individuals that are currently infectious and does not account for individuals that have already recovered. Finally, the ratio of secondary case production caused by asymptomatic vs. symptomatic individuals during the exponential phase should be

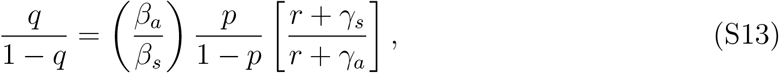

where *q* is the fraction of new secondary cases caused by asymptomatic individuals.

The basic reproduction number of this system is:

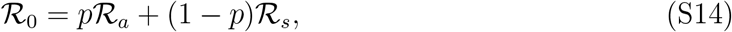

where

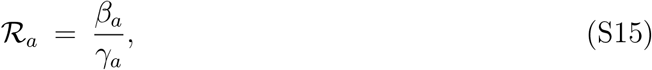

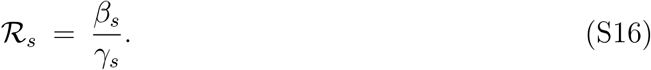

The generation-interval distributions for asymptomatic and symptomatic individuals follow the same functional form as the corresponding generation-interval distribution for a single-type SEIR model since both asymptomatic and symptomatic individuals have exponentially distributed latent and infectious periods [25]:

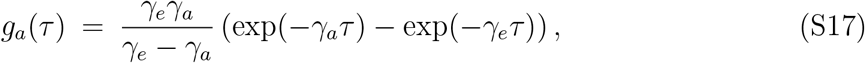

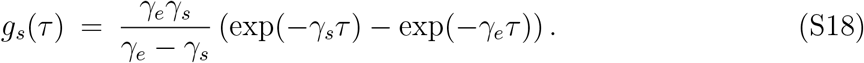

It immediately follows that

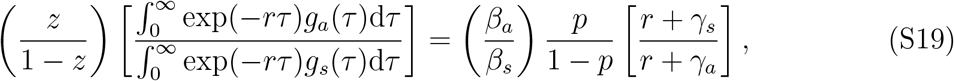

where *z* = *p ℛ*_*a*_*/ℛ*_0_ and 1 *− z* = (1 *− p*)*ℛ*_*s*_*/ℛ*_0_ – compare to Eq. (8) in the main text.

## Supplementary table

**TABLE S1:**
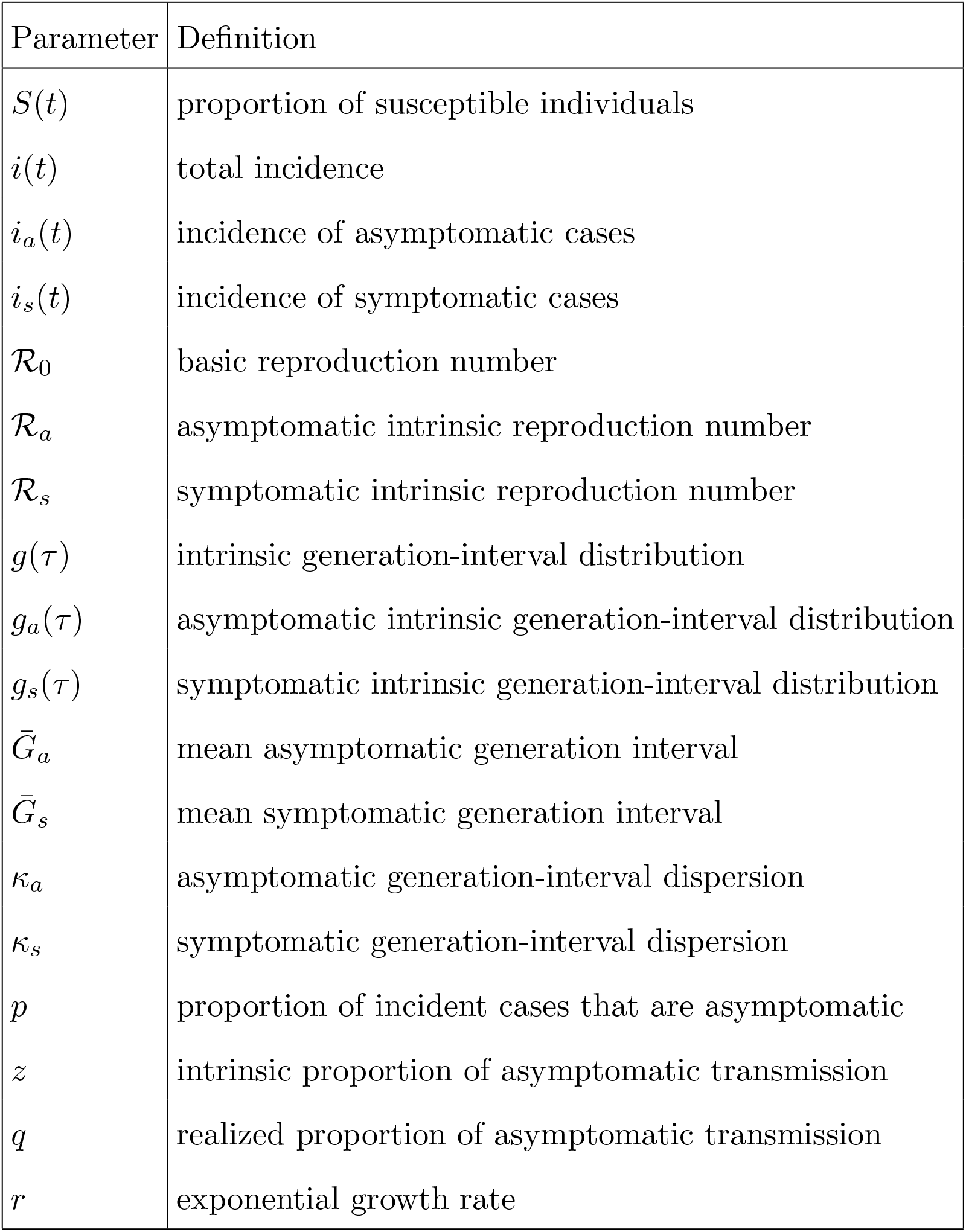
Model parameters and definitions.

## Supplementary figures

**FIG. S1:**
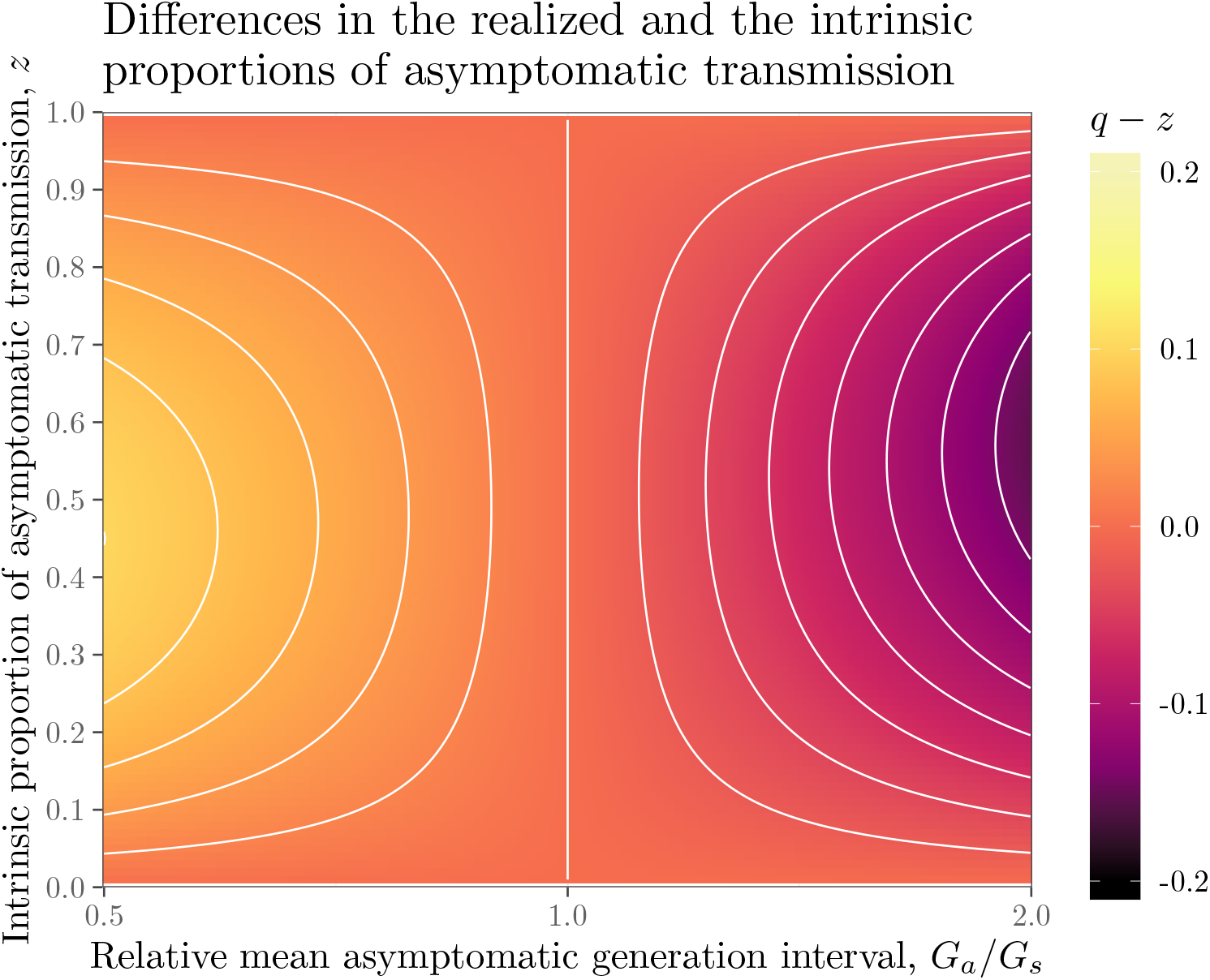
Differences in the realized and the intrinsic proportions of asymptomatic transmission. See Figure 1 in the main text for figure caption.

**FIG. S2:**
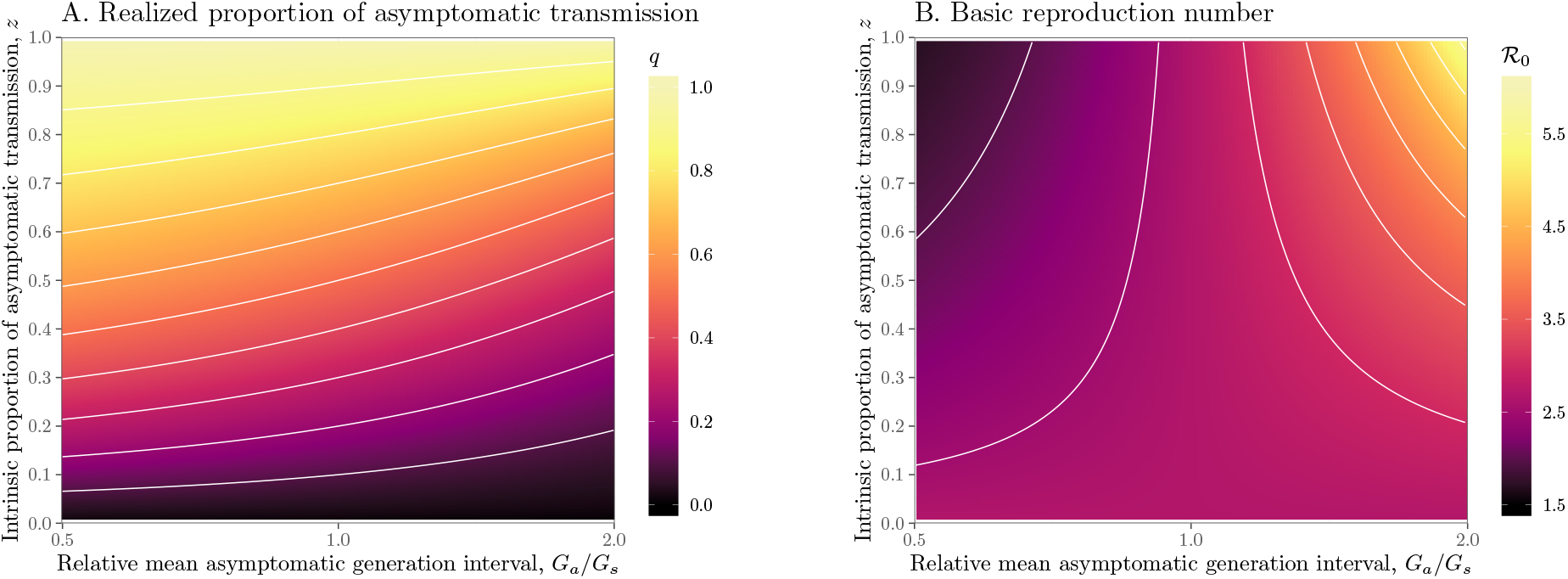
Effects of intrinsic proportion of asymptomatic transmission on the realized proportion of asymptomatic transmission and basic reproduction number, given variation in the mean generation interval of asymptomatic cases when generation-interval distributions are narrow. Here, we assume 1*/r* = 7 days, 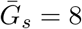 = 8 days, and *κ*_*s*_ = *κ*_*a*_ = 0.3. See Figure 1 in the main text for figure caption.

**FIG. S3:**
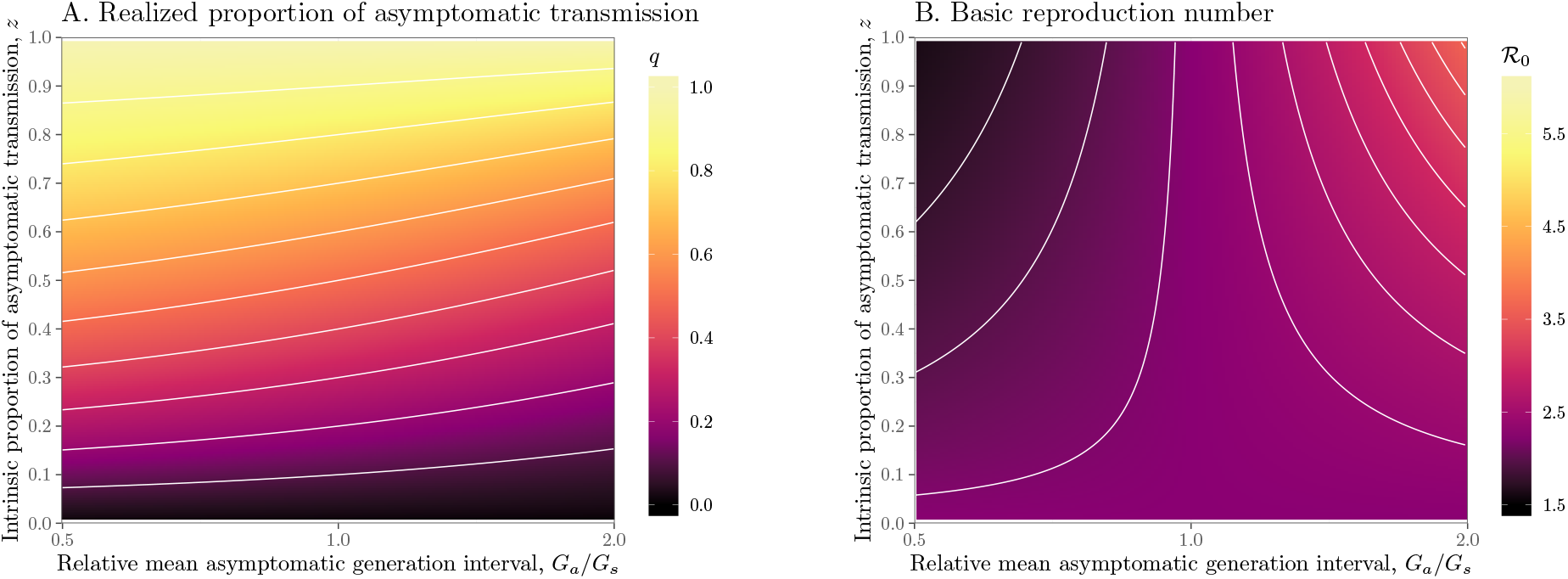
Effects of intrinsic proportion of asymptomatic transmission on the realized proportion of asymptomatic transmission and basic reproduction number, given variation in the mean generation interval of asymptomatic cases when generation-interval distributions are wide. Here, we assume 1*/r* = 7 days, 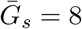 days, and *κ*_*s*_ = *κ*_*a*_ = 0.8. See Figure 1 in the main text for figure caption.

**FIG. S4:**
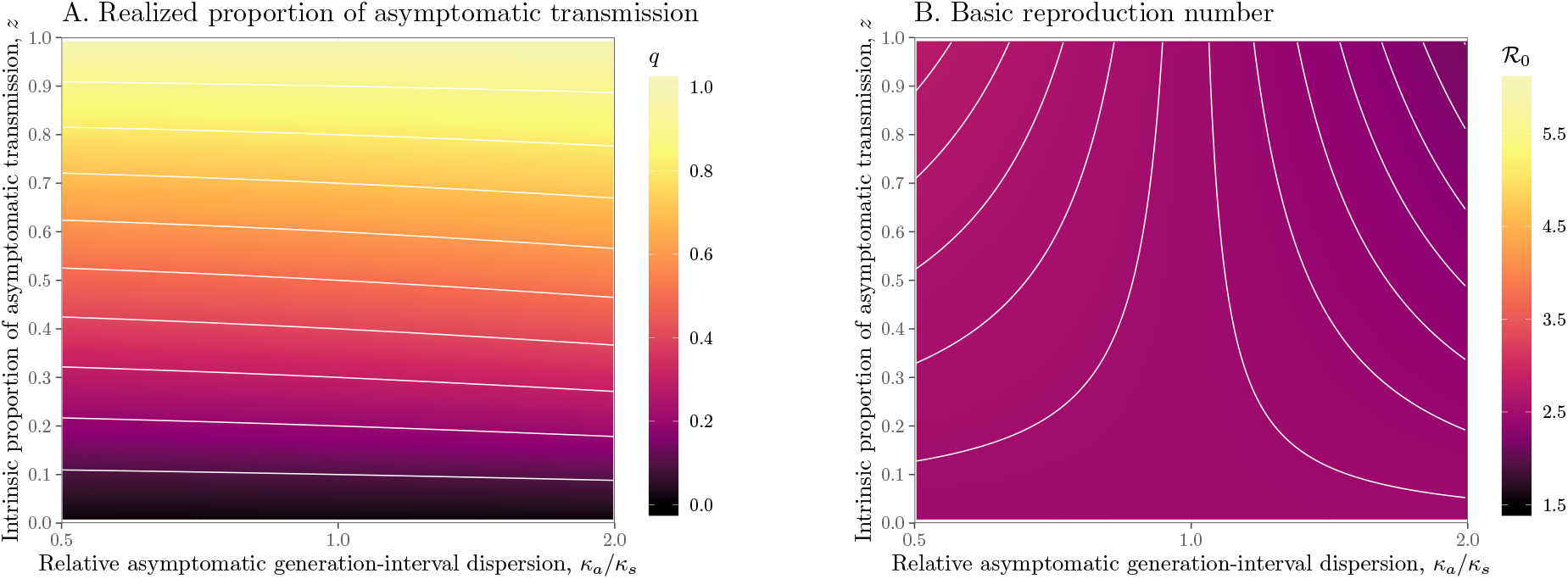
Effects of intrinsic proportion of asymptomatic transmission on the realized proportion of asymptomatic transmission and basic reproduction number, given variation in the generation-interval dispersion of asymptomatic cases. (A) Wide/narrow generation intervals of asymptomatic cases increase/decrease the relevance of asymptomatic cases, *q*. (B) Wide/narrow generation intervals of asymptomatic cases decrease/increase the basic reproduction number *ℛ*_0_. Solid lines show contours for *q* and *ℛ*_0_ values. Here, we assume 1*/r* = 7 days, 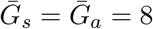 days, and *κ*_*s*_ = 0.5.

## References

[1] Fraser, C., Riley, S., Anderson, R. M. & Ferguson, N. M. Factors that make an infectious disease outbreak controllable. Proceedings of the National Academy of Sciences 101, 6146–6151 (2004).

[2] Quilty, B. J., Clifford, S., CMMID nCoV working group2, Flasche, S. & Eggo, R. M. Effectiveness of airport screening at detecting travellers infected with novel coronavirus (2019-nCoV). Eurosurveillance 25 (2020).

[3] Chan, J. F.-W. et al. A familial cluster of pneumonia associated with the 2019 novel coronavirus indicating person-to-person transmission: a study of a family cluster. The Lancet 395, 514–523 (2020).

[4] Pan, X. et al. Asymptomatic cases in a family cluster with SARS-CoV-2 infection. The Lancet Infectious Diseases (2020).

[5] Tang, A. et al. Detection of novel coronavirus by RT-PCR in stool specimen from asymptomatic child, China. Emerging infectious diseases 26 (2020).

[6] Bai, Y. et al. Presumed asymptomatic carrier transmission of COVID-19. JAMA (2020).

[7] Mizumoto, K., Kagaya, K., Zarebski, A. & Chowell, G. Estimating the asymptomatic proportion of coronavirus disease 2019 (COVID-19) cases on board the Diamond Princess cruise ship, Yokohama, Japan, 2020. Eurosurveillance 25 (2020).

[8] Fauci, A. S., Lane, H. C. & Redfield, R. R. Covid-19 – navigating the uncharted. New England Journal of Medicine (2020). URL https://doi.org/10.1056/NEJMe2002387.

[9] Li, R. et al. Substantial undocumented infection facilitates the rapid dissemination of novel coronavirus (sars-cov2). Science (2020). URL https://science.sciencemag.org/content/early/2020/03/13/science.abb3221. https://science.sciencemag.org/content/early/2020/03/13/science.abb3221.full.pdf.

[10] Anderson, R. M. & May, R. M. Infectious diseases of humans: dynamics and control (Oxford university press, 1992).

[11] Heesterbeek, J. & Dietz, K. The concept of R_0_ in epidemic theory. Statistica Neerlandica 50, 89–110 (1996).

[12] Champredon, D. & Dushoff, J. Intrinsic and realized generation intervals in infectious-disease transmission. Proceedings of the Royal Society B: Biological Sciences 282, 20152026 (2015).

[13] Svensson, °A. A note on generation times in epidemic models. Mathematical Biosciences 208, 300–311 (2007).

[14] Wallinga, J. & Lipsitch, M. How generation intervals shape the relationship between growth rates and reproductive numbers. Proceedings of the Royal Society B: Biological Sciences 274, 599–604 (2007).

[15] Roberts, M. & Heesterbeek, J. Model-consistent estimation of the basic reproduction number from the incidence of an emerging infection. Journal of Mathematical Biology 55, 803 (2007).

[16] Park, S. W. et al. Reconciling early-outbreak estimates of the basic reproductive number and its uncertainty: framework and applications to the novel coronavirus (SARS-CoV-2) outbreak. medRxiv (2020). URL https://www.medrxiv.org/content/10.1101/2020.01.30.20019877v4.full.pdf.

[17] Lotka, A. J. Relation between birth rates and death rates. Science 26, 21–22 (1907).

[18] Park, S. W., Champredon, D., Weitz, J. S. & Dushoff, J. A practical generation-interval-based approach to inferring the strength of epidemics from their speed. Epidemics 27, 12–18 (2019).

[19] Wearing, H. J., Rohani, P. & Keeling, M. J. Appropriate models for the management of infectious diseases. PLoS Medicine 2 (2005).

[20] Powers, K. A., Kretzschmar, M. E., Miller, W. C. & Cohen, M. S. Impact of early-stage HIV transmission on treatment as prevention. Proceedings of the National Academy of Sciences 111, 15867–15868 (2014).

[21] Watts, D. J., Muhamad, R., Medina, D. C. & Dodds, P. S. Multiscale, resurgent epidemics in a hierarchical metapopulation model. Proceedings of the National Academy of Sciences 102, 11157–11162 (2005).

[22] Chinazzi, M. et al. The effect of travel restrictions on the spread of the 2019 novel coronavirus (COVID-19) outbreak. Science (2020).

[23] Du, Z. et al. Risk for transportation of 2019 novel coronavirus disease from Wuhan to other cities in China. Emerging Infectious Diseases 26 (2020).

[24] Kissler, S. M., Tedijanto, C., Goldstein, E., Grad, Y. H. & Lipsitch, M. Projecting the transmission dynamics of SARS-CoV-2 through the post-pandemic period. medRxiv (2020). URL https://www.medrxiv.org/content/10.1101/2020.03.04.20031112v1.

[25] Svensson, °A. The influence of assumptions on generation time distributions in epidemic models. Mathematical Biosciences 270, 81–89 (2015).

